# Substantial undocumented infection facilitates the rapid dissemination of novel coronavirus (COVID-19)

**DOI:** 10.1101/2020.02.14.20023127

**Authors:** Ruiyun Li, Sen Pei, Bin Chen, Yimeng Song, Tao Zhang, Wan Yang, Jeffrey Shaman

**Author notes:** Correspondence to: S.P. J.S. R.L., S.P. and B.C. contributed equally to this work.

## Abstract

**Background:** Estimation of the fraction and contagiousness of undocumented novel coronavirus (COVID-19) infections is critical for understanding the overall prevalence and pandemic potential of this disease. Many mild infections are typically not reported and, depending on their contagiousness, may support stealth transmission and the spread of documented infection.

**Methods:** Here we use observations of reported infection and spread within China in conjunction with mobility data, a networked dynamic metapopulation model and Bayesian inference, to infer critical epidemiological characteristics associated with the emerging coronavirus, including the fraction of undocumented infections and their contagiousness.

**Results:** We estimate 86% of all infections were undocumented (95% CI: [82%-90%]) prior to the Wuhan travel shutdown (January 23, 2020). Per person, these undocumented infections were 52% as contagious as documented infections ([44%-69%]) and were the source of infection for two-thirds of documented cases. Our estimate of the reproductive number (2.23; [1.77-3.00]) aligns with earlier findings; however, after travel restrictions and control measures were imposed this number falls considerably.

**Conclusions:** A majority of COVID-19 infections were undocumented prior to implementation of control measures on January 23, and these undocumented infections substantially contributed to virus transmission. These findings explain the rapid geographic spread of COVID-19 and indicate containment of this virus will be particularly challenging. Our findings also indicate that heightened awareness of the outbreak, increased use of personal protective measures, and travel restriction have been associated with reductions of the overall force of infection; however, it is unclear whether this reduction will be sufficient to stem the virus spread.

---

The novel coronavirus that emerged in Wuhan, China (COVID-19) at the end of 2019 quickly spread to all Chinese provinces and, as of February 6, 2020, to 24 other countries^1,2^. Efforts to contain the virus are ongoing; however, given the many uncertainties regarding pathogen transmissibility and virulence, the effectiveness of these efforts is unknown.

The fraction of undocumented but infectious cases is a critical epidemiological characteristic that modulates the pandemic potential of an emergent respiratory virus^3–6^. These undocumented infections often experience mild, limited or no symptoms and hence go unrecognised, and, depending on their contagiousness and numbers, can expose a far greater portion of the population to virus than would otherwise occur. Here, to assess the full potential of COVID-19, we use a model-inference framework to estimate the contagiousness and proportion of undocumented infections in China during the weeks before and after the shutdown of travel in and out of Wuhan.

## Methods

### Metapopulation Model

We developed a mathematical model that simulates the spatio-temporal dynamics of infections among 375 Chinese cities. The model incorporates information on human movement within the following metapopulation structure:

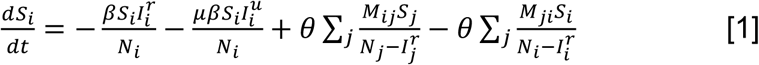

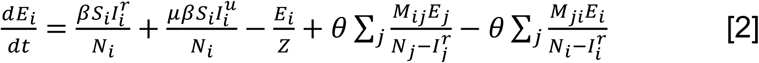

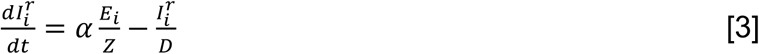

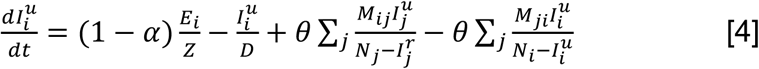

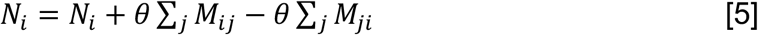

where 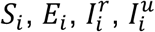 and *N*_*i*_ are the susceptible, exposed, documented infected, undocumented infected and total population in city *i*. Note that we define patients with symptoms severe enough to be confirmed as documented infected individuals; whereas other infected persons are defined as undocumented infected individuals. We specified a rate parameter, *β*, for the transmission rate due to documented infected individuals. The transmission rate due to undocumented individuals is reduced by a factor *μ*. In addition, *α* is the fraction of documented infections, *Z* is the average latency period and *D* is the average duration of infection. The effective reproduction number (*R*_*E*_) is calculated as *R*_*E*_ = *αβD* + (1 − *α*)*μβD* (see Supplementary Appendix for details). Spatial coupling within the model is represented by the daily number of people traveling from city *j* to city *i* (*M*_*ij*_) and a multiplicative factor, *θ*, which is greater than 1 to reflect underreporting of human movements (see below). We assume that individuals in the 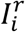 group do not move between cities. A similar metapopulation model has been used to forecast the spatial transmission of influenza in the United States^7^.

### Travel Data

Daily numbers of travelers between 375 Chinese cities during the Spring Festival period (“Chunyun”) were derived from human mobility data collected by the Tencent Location-based Service (LBS) during the 2018 Chunyun period (February 1 – March 12, 2018) ^8^. Chunyun is a period of 40 days – 15 days before and 25 days after the Lunar New Year – during which there are high rates of travel within China. To estimate human mobility during the 2020 Chunyun period, which began January 10, we aligned the 2018 Tencent data based on relative timing to the Spring Festival. For example, we used mobility data from February 1, 2018 to represent human movement on January 10, 2020, as these days were similarly distant from the Lunar New Year. During the 2018 Chunyun, a total of 1.73 billion travel events were captured in the Tencent data; whereas 2.97 billions trips are reported^8^. To reconcile these two numbers, we include the parameter *θ* in the model system.

### Inference and Model Initialization

To infer COVID-19 transmission dynamics during the early stage of the outbreak, we simulated observations from January 10-23, 2020 (i.e. the period before the initiation of travel restrictions) using an iterated filter-ensemble adjustment Kalman filter (IF-EAKF) framework^9-11^. With this combined model-inference system, we estimated the trajectories of the four model state variables 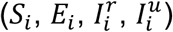 for all 375 cities, while simultaneously inferring the six model parameters (*Z, D, μ, β, α, θ*). The initial prior ranges of the model parameters were drawn from uniform distributions of the following ranges: 2 *days* ≤ *Z* ≤ 5 *days*, 2 *days* ≤ *D* ≤ 5 *days*, 0.2 ≤ *μ* ≤ 1, 0.6 ≤ *β* ≤ 1.5, 0.02 ≤ *α* ≤ 0.8, 1 ≤ *θ* ≤ 1.75.

For the outbreak origin, Wuhan city, the initial exposed population, *E*_*wuhan*_, and initial undocumented infected population, 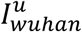, were drawn from a uniform distribution [0, *Seed*_*max*_]. The documented infected population in Wuhan 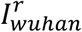 on January 10 was set to zero. Although infections were reported prior to January 10, these cases were sporadic and the EAKF adjustment can account for the effects of these early infections (by selecting elevated exposed and unreported infection levels). For other cities, we defined *C*_*i*_ as the number of travelers from Wuhan to city *i* on the first day of Chunyun. The initial exposed, documented infected and undocumented infected populations were set to 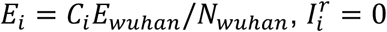 and 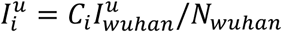.

To account for delays in infection confirmation, we also defined an observation model using a Poisson process. Specifically, for each new case in group 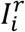, a reporting delay *t*_*d*_ (in days) was generated from a Poisson distribution with a mean value of *T*_*d*_. In fitting both synthetic and the observed outbreaks, we performed simulations with the model-inference system using different fixed values of *T*_*d*_ (4 *days* ≤ *T*_*d*_ ≤ 12 *days*) and *Seed*_*max*_ (500 ≤ *Seed*_*max*_ ≤ 6000). The best fitting model-inference posterior was identified by log-likelihood. Full details of the data and methods, including synthetic testing and sensitivity analyses, are provided in the Supplementary Appendix.

### Modelling epidemic dynamics after January 23

Finally, we also modelled the transmission of COVID-19 in China after January 23, when greater control measures were effected. These control measures included travel restrictions imposed between major cities and Wuhan; self-quarantine and contact precautions advocated by the government; and more available rapid testing for infection confirmation^12-13^. These measures along with changes in medical care-seeking behaviour due to increased awareness of the virus and increased personal protective behavior (e.g. wearing of facemasks, social distancing, self-isolation when sick), likely altered the epidemiological characteristics of the outbreak after January 23. To quantify these differences, we re-estimated the system parameters using the metapopulation model-inference framework and city-level daily cases reported between January 24 and February 8. As inter-city mobility was restricted, we set *θ* = 0. In addition, to represent reduced person-to-person contact and increased infection detection, we updated the initial priors for *β* and *α* to [0.2, 1.0] and [0.2, 1.0], respectively (see Supplementary Appendix for more details).

## Results

### Epidemiological Characteristics before January 23, 2020

We first tested the model-inference framework using synthetic outbreaks generated by the model in free simulation. These simulations verified the ability of the model-inference framework to simultaneously estimate the six target model parameters (see Supplementary Appendix, Figures S1-S8).

We next applied the system to the observed outbreak before the travel restrictions of January 23 – a total of 811 documented cases throughout China. Figure 1 shows simulations of reported cases generated using the best-fitting model parameter estimates. The distribution of these stochastic simulations captures the range of observed cases well. In addition, the best-fitting model captures the spread of COVID-19 to other cities in China (Figure S9). Our median estimate of the overall *R*_*E*_ is 2.23 (95% CI: 1.77−3.00), indicating a high capacity for sustained transmission of COVID-19 (Table 1). This finding aligns with other recent estimates of the reproductive number for this time period^6,12-14^. In addition, the median estimates for the latent and infectious periods are approximately 3.77 and 3.45 days, respectively. Further, we find that, during January 10-23, only 14% (95% CI: 9–26%) of total infections in China were reported. This estimate reveals a very high rate of undocumented infections: 86%. This finding is independently corroborated by the infection rate among foreign nationals evacuated from Wuhan (see Supplementary Appendix). These undocumented infections are estimated to have been half as contagious per individual as reported infections (*µ* = 0.52; 95% CI: 0.44 – 0.69). Other model fittings made using alternate values of *T*_*d*_ and *Seed*_*max*_ produced similar parameter estimates (Figure S10).

**Table 1.**
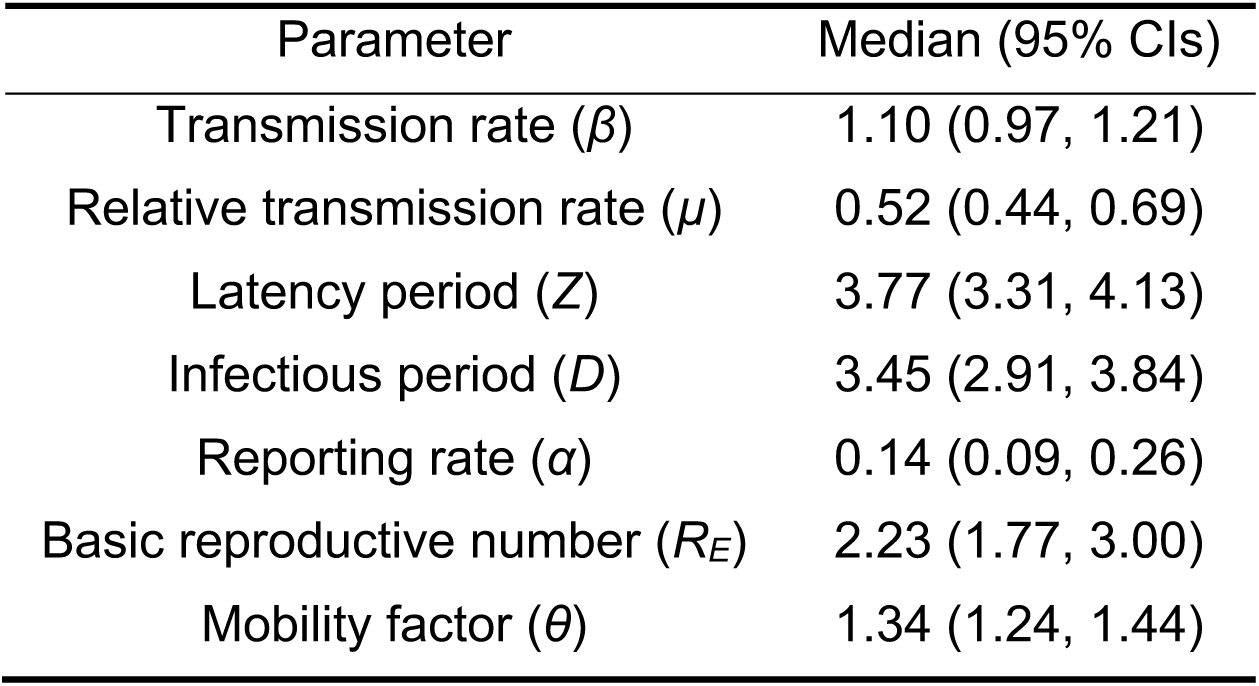
Best-fit model posterior estimates of key epidemiological parameters for simulation with the full metapopulation model during January 10-23, 2020 (*Seed*_*max*_ = 5000, *T*_*d*_ = 10 days).

| Parameter | Median (95% CIs) |
| --- | --- |
| Transmission rate ($\beta$) | 1.10 (0.97, 1.21) |
| Relative transmission rate ($\mu$) | 0.52 (0.44, 0.69) |
| Latency period ($Z$) | 3.77 (3.31, 4.13) |
| Infectious period ($D$) | 3.45 (2.91, 3.84) |
| Reporting rate ($\alpha$) | 0.14 (0.09, 0.26) |
| Basic reproductive number ($R_E$) | 2.23 (1.77, 3.00) |
| Mobility factor ($\theta$) | 1.34 (1.24, 1.44) |

**Fig. 1.**
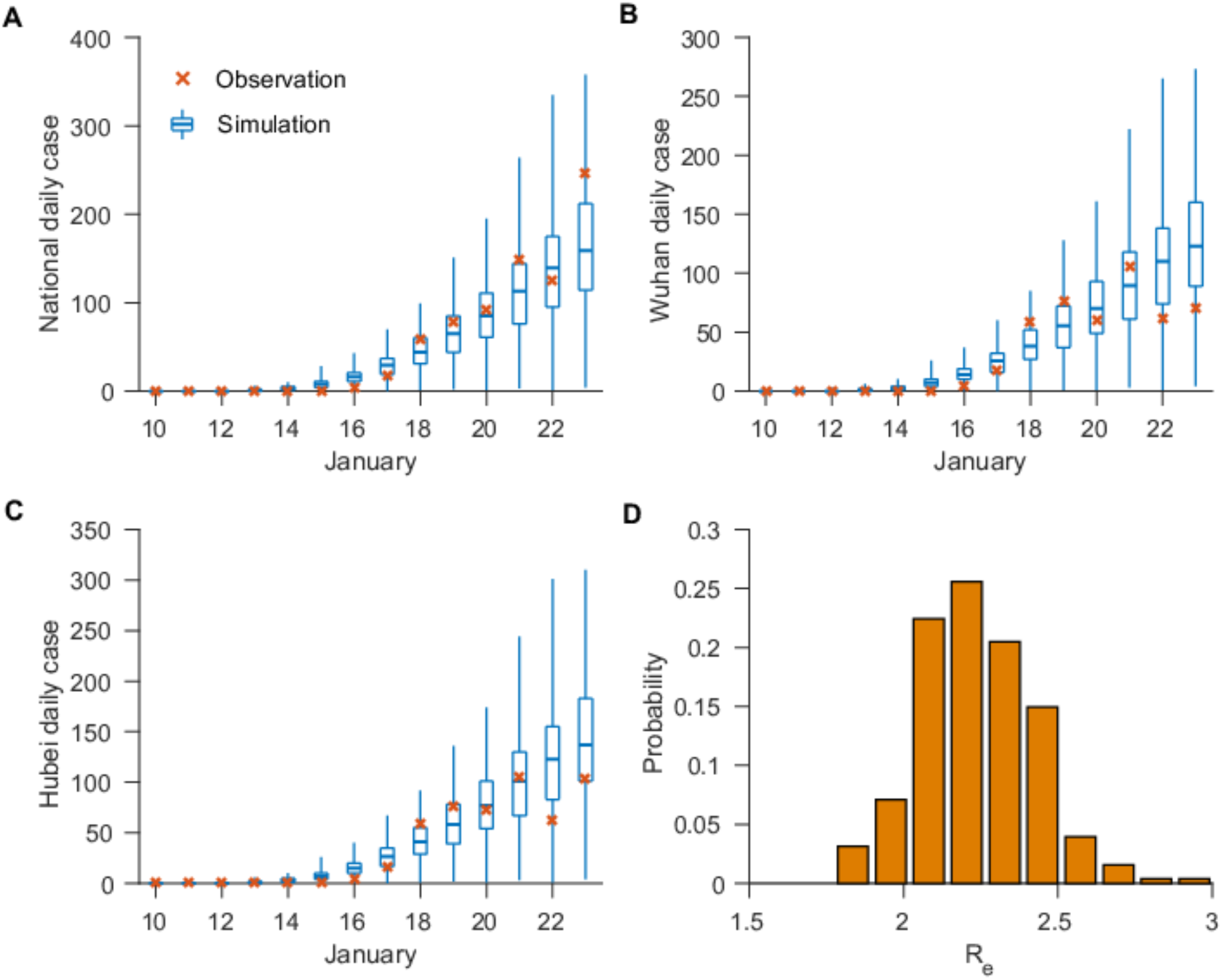
Best-fit model-inference fitting (*Seed*_*max*_ = 5000, *T*_*d*_ = 10 days) to daily reported cases in all cities (A), Wuhan city (B) and Hubei province (C). The blue box and whiskers show the median, interquartial range, and 95% credible intervals are derived from 300 simulations using the best-fit parameters. The red ‘x’s are daily reported cases. The distribution of estimated *R*_*E*_ is shown in (D).

### The Impact of Undocumented Infections during January 10-23

Using the best-fitting model (Table 1, Figure 1), we estimated 18,829 (95% CI [3,761, 38,808]) total new COVID-19 infections (documented and undocumented combined) during January 10-23 in Wuhan city. 86.3% of all infections (95% CI [81.9%, 90.1%]) were infected from undocumented cases. Nationwide, the total number of infections during January 10-23 was 28,898 (95% CI [5,534, 59,491]) with 86.4% (95% CI [82.0%, 90.1%]) infected from undocumented cases.

To highlight further this impact of contagious, undocumented COVID-19 infections on overall transmission and reported case counts, we generated a set of hypothetical outbreaks using the best-fitting parameter estimates but with *μ* = 0, i.e. the undocumented infections are no longer contagious (Figure 2). We find that without transmission from undocumented cases, reported infections during January 10-23 are reduced 66.4% across all of China and 64.0% in Wuhan. Further, there are fewer cities with more than 8 cumulative documented cases: only 1 city with more than 8 documented cases versus the 10 observed by January 23 (Figure 2). This finding indicates that contagious, undocumented infections facilitated the geographic spread of COVID-19 within China.

**Fig. 2.**
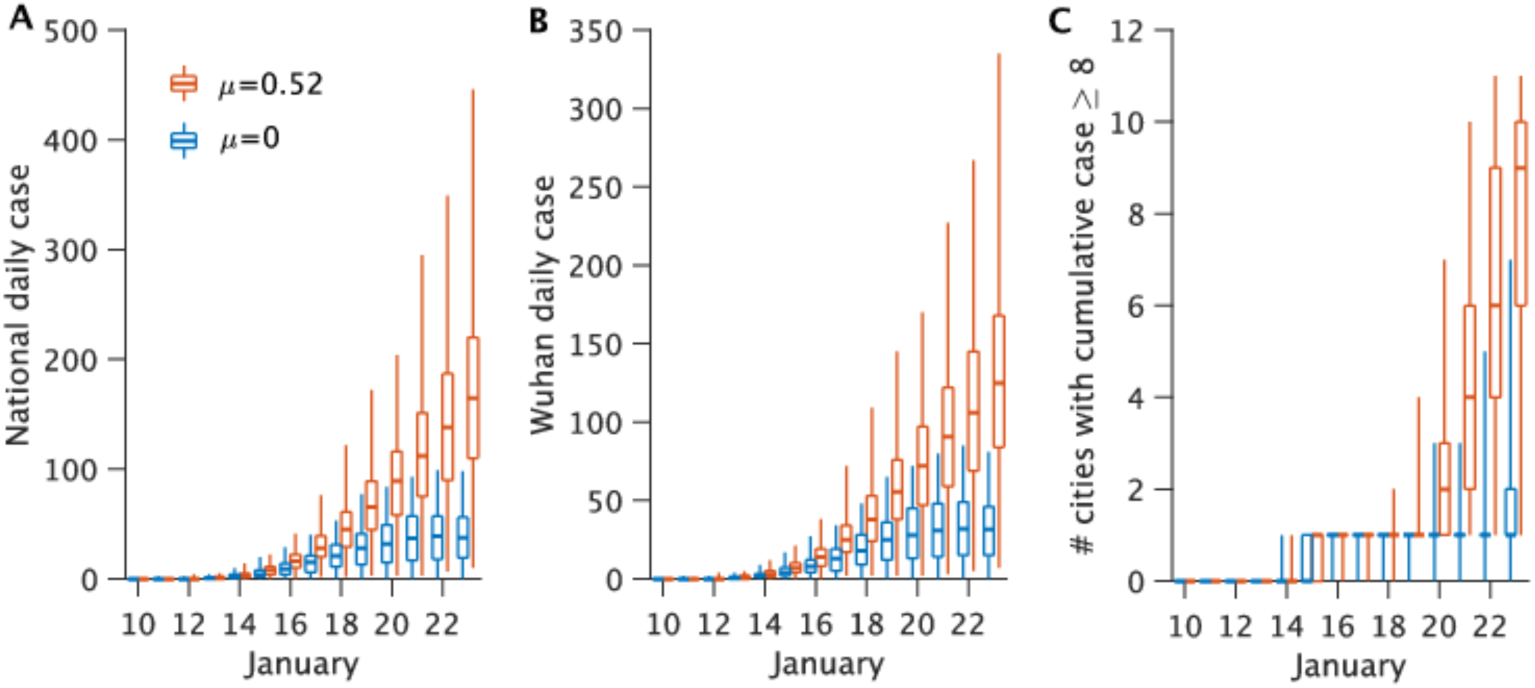
Impact of undocumented infections on the transmission of COVID-19. Synthetic outbreaks generated using parameters reported in Table 1 are compared for *μ* = 0.52 (red) and *μ* = 0 (blue).

### Epidemiological Characteristics after January 23, 2020

The results of inference for the January 24-February 8 period are presented in Table 2, Figure S11 and Table S1. Control measures are continually shifting, so we show estimates for both January 24 – February 3 (Period 1) and January 24 – February 8 (Period 2). The best-fitting model for both periods has a reduced reporting delay, *T*_*d*_, of 5 days (vs. 10 days before January 23), consistent with more rapid confirmation of infections. Estimates of both the latency and infectious periods are relatively unchanged; however, *α, β* and *R*_*E*_ have all shifted considerably. The contact rate, *β*, drops to 0.51 (95% CI: 0.39 – 0.69) during Period 1 and 0.34 (95% CI: 0.27 – 0.48) during Period 2, less than half the estimate prior to travel restrictions. The reporting rate, *α*, is estimated to be 0.71 (95% CI: 0.55 – 0.85), i.e. 71% of infections are documented during Period 1, up from 0.14 prior to travel restrictions, and is nearly the same in Period 2. The reproductive number is 1.51 (95% CI: 1.17 – 2.10) during Period 1 and 1.00 (95% CI: 0.73 – 1.38) during Period 2, down from 2.23 prior to travel restrictions. While the estimate for the relative transmission rate, *μ*, is similar to before January 23, the contagiousness of undocumented infections, represented by *μβ*, is substantially reduced, possibly reflecting that only very mild and asymptomatic infections remain undocumented.

**Table 2.** Best-fit model posterior estimates of key epidemiological parameters for simulation of the model without travel between cities during January 24 – February 3 and January 24 – February 8 (*Seed*_*max*_ = 5000 on January 10, *T*_*d*_ = 10 days before January 24, *T*_*d*_ = 5 days between January 24 and February 8).

| Parameter | January 24 – February 3<br>(Median (95% CIs)) | January 24 - February 8<br>(Median (95% CIs)) |
| --- | --- | --- |
| Transmission rate ($\beta$) | 0.51 (0.39, 0.69) | 0.34 (0.27, 0.48) |
| Relative transmission rate ($\mu$) | 0.49 (0.38, 0.60) | 0.43 (0.29, 0.67) |
| Latency period ($Z$) | 3.49 (3.35, 3.68) | 3.50 (3.23, 3.77) |
| Infectious period ($D$) | 3.50 (3.28, 3.64) | 3.51 (3.19, 3.82) |
| Reporting rate ($\alpha$) | 0.71 (0.56, 0.81) | 0.71 (0.56, 0.85) |
| Effective reproductive number<br>($R_E$) | 1.51 (1.17, 2.10) | 1.00 (0.73, 1.38) |

## Discussion

Our findings indicate that a large proportion of COVID-19 infections were undocumented prior to the implementation of travel restrictions and other heightened control measures in China on January 23, and that a large proportion of the total force of infection was mediated through these undocumented infections (Table 1). This high proportion of undocumented infections, many of whom were likely not severely symptomatic, appears to have supported the rapid spread of the virus throughout China. Indeed, suppression of the infectiousness of these undocumented cases in model simulations reduces the total number of documented cases and the overall spread of COVID-19 (Figure 2).

Our findings also indicate that a radical increase in the identification and isolation of currently undocumented infections would be needed to fully control COVID-19.

Increased news coverage and awareness of the virus in the general population have already likely prompted increased rates of seeking medical care for respiratory symptoms. In addition, awareness among healthcare providers, public health officials and the availability of viral identification assays suggest that capacity for identifying previously missed infections has increased. Further, general population and government response efforts have increased the use of face masks, restricted travel, delayed school reopening and isolated suspected persons, all of which could additionally slow the spread of COVID-19.

Combined, these measures are expected to increase reporting rates, reduce the proportion of undocumented infections, and decrease the growth and spread of infection. Indeed, estimation of the epidemiological characteristics of the outbreak after January 23, indicate that government control efforts and population awareness have reduced the rate of spread of the virus (i.e. lower *β, μβ, R*_*E*_) and increased the reporting rate. The overall reduction of the effective reproductive number is encouraging; however, the control efforts have yet to critically and clearly reduce *R*_*E*_ below 1.

Importantly, the situation on the ground in China is changing day-to-day. New travel restrictions and control measures are being imposed on new populations in different cities, and these rapidly varying effects make certain estimation of the epidemiological characteristics for the outbreak difficult. Further, reporting inaccuracies and changing care-seeking behavior add another level of uncertainty to our estimations. While the data and findings presented here indicate that travel restrictions and control measures have reduced COVID-19 transmission considerably, whether these controls are sufficient for reducing *R*_*E*_ below 1 for the length of time needed to eliminate the disease locally and prevent a rebound outbreak once control measures are relaxed is unclear. Further, similar control measures and travel restrictions would have to be implemented outside China to prevent re-introduction of the virus.

Our findings underscore the seriousness and pandemic potential of COVID-19. The 2009 H1N1 pandemic influenza virus also caused many mild cases, quickly spread globally, and eventually became endemic. Presently, there are four, endemic, coronavirus strains currently circulating in human populations (229E, HKU1, NL63, OC43). If the novel coronavirus follows the pattern of 2009 H1N1 pandemic influenza, it will also spread globally and become a fifth endemic coronavirus within the human population.

Many characteristics of the COVID-19 remain unknown or uncertain. Consequently, care should be taken when interpreting our estimates. For instance, after January 23, we assume a complete travel shutdown with no inter-city human mobility; however, the degree and initial date of travel restrictions has varied among cities. Our estimates may therefore represent an upper-bound of the potential impact of travel restriction on COVID-19 transmission. Further studies accounting for heterogenous travel interventions are warranted.

## Data Availability

All data are publicly available.

## Funding

This work was supported by US NIH grants GM110748 and AI145883. The content is solely the responsibility of the authors and does not necessarily represent the official views of the National Institute of General Medical Sciences, the National Institute for Allergy and Infectious Diseases, or the National Institutes of Health.

## Disclosures

JS and Columbia University disclose partial ownership of SK Analytics. JS also reports receiving consulting fees from Merck.

